# Epistatic Features and Machine Learning Improve Alzheimer’s Risk Prediction Over Polygenic Risk Scores

**DOI:** 10.1101/2023.02.10.23285766

**Authors:** Stephen Hermes, Janet Cady, Steven Armentrout, James O’Connor, Sarah Carlson, Carlos Cruchaga, Thomas Wingo, Ellen McRae Greytak, The Alzheimer’s Disease Neuroimaging Initiative

## Abstract

**Background:** Polygenic risk scores (PRS) are linear combinations of genetic markers weighted by effect size that are commonly used to predict disease risk. For complex heritable diseases such as late onset Alzheimer’s disease (LOAD), PRS models fail to capture much of the heritability. Additionally, PRS models are highly dependent on the population structure of data on which effect sizes are assessed, and have poor generalizability to new data.

**Objective:** The goal of this study is to construct a paragenic risk score that, in addition to single genetic marker data used in PRS, incorporates epistatic interaction features and machine learning methods to predict lifetime risk for LOAD.

**Methods:** We construct a new state-of-the-art genetic model for lifetime risk of Alzheimer’s disease. Our approach innovates over PRS models in two ways: First, by directly incorporating epistatic interactions between SNP loci using an evolutionary algorithm guided by shared pathway information; and second, by estimating risk via an ensemble of machine learning models (gradient boosting machines and deep learning) instead of simple logistic regression. We compare the paragenic model to a PRS model from the literature trained on the same dataset.

**Results:** The paragenic model is significantly more accurate than the PRS model under 10-fold cross-validation, obtaining an AUC of 83% and near-clinically significant matched sensitivity/specificity of 75%, and remains significantly more accurate when evaluated on an independent holdout dataset. Additionally, the paragenic model maintains accuracy within APOE genotypes.

**Conclusion:** Paragenic models show potential for improving lifetime disease risk prediction for complex heritable diseases such as LOAD over PRS models.

## 1. Introduction

Alzheimer’s Disease (AD) is the most common cause of dementia, affecting millions of Americans, and is the only disease among the leading causes of death in the US for which no effective prevention or cure exists [1]. The FDA recently drafted a set of industry guidelines for clinical trials of AD treatments targeting the earliest stages of disease [2], indicating increasing focus on and investment in presymptomatic intervention. However, trials aimed at averting the underlying causes of disease have proven difficult because pathological changes in AD happen well in advance of cognitive decline. While changes in levels of amyloid-/3 (A/3) and tau in cerebrospinal fluid [3] and even blood [4] can be seen prior to onset of symptoms, these changes indicate that pathogenic processes have already begun. Furthermore, a biomarker test administered too far in advance of symptom onset may not indicate future risk of developing AD. An accurate genetic test for AD, on the other hand, could be used at any point in life to identify individuals at high risk for developing the disease before changes in biomarkers can be detected.

Development of such a test is complicated by the complex genetic structure of the more common, late-onset form of AD (LOAD). The strongest risk factor for LOAD, the Apolipoprotein E (*APOE*) *ε*4 allele, increases risk of developing LOAD ≈15 fold for those with two copies of the *ε*4 allele and ≈ 3 fold for those with one copy [5]; however, it only accounts for ≈6% of phenotypic variance [6]. Many additional genetic risk factors have since been identified, all having much smaller effect sizes.

Recent genetic risk prediction models for LOAD have attempted to capture this complexity using polygenic risk scores (PRS), in which an individual’s risk is calculated by summing their total number of risk alleles across multiple markers, weighted by effect size. PRS models for LOAD have reported area under the receiver operator characteristic curve (AUC) ranging from 0.62–0.78 for clinically diagnosed LOAD [7] and 0.82 in pathologically confirmed cases [8]. These models focus only on the additive effects of single nucleotide polymorphisms (SNPs) leaving a significant amount of heritability unexplained; of the nearly 75% estimated heritability of LOAD [9], only 24% is explained by additive genetic components [10]. PRS models capture only 21% of the estimated heritability (90% of the heritability explained by additive genetic components) [8].

One possible source of missing heritability is nonadditive, or epistatic, interactions between SNPs. Epistatic interactions have been discovered involving genes that are independently associated with LOAD, as well as between genes that are not significantly associated with LOAD on their own [11]. A recent study [12] constructed a LOAD genetic risk prediction model combining epistatic risk with polygenic risk and achieved an AUC of 0.67. Although this was lower than other reported PRS models, it was a minor improvement over their model using only PRS scores in the same dataset. Genome-wide epistasis studies are often limited to two-way interactions between SNPs; the large number of SNPs means that the number of possible genotype combinations for higher order interactions is virtually infinite.

In this paper, we use Crush-MDR [13], a machine learning algorithm that combines multifactor dimensionality reduction (MDR) with an evolutionary search algorithm, to identify epistatic interactions in LOAD. These interactions are included with single SNPs and PRS values to produce a state-of-the-art LOAD risk prediction model. We term our model a *paragenic risk model* as it incorporates genetic markers beyond individual SNPs and it is an ensemble consisting of a PRS model along with machine learning models. The paragenic risk model shows significant improvement over PRS and gradient boosting machines alone, obtaining a mean 10-fold cross-validated area under the receiver operator characteristic curve (AUC) of 0.83 (95% CI [0.82, 0.84]) in predicting LOAD in clinically diagnosed cases. Additionally, our paragenic model maintains high AUC within *APOE* genotype strata, unlike PRS models.

## 2. Methods

### 2.1. Participants

The dataset used for modeling consisted of data from the Alzheimer’s Disease Neuroimaging Initiative (ADNI), the National Alzheimer’s Coordinating Center and the Alzheimer’s Disease Genetics Consortium (NACC/ADGC), the Framingham Heart Study (FHS), the Knight-ADRC at Washington University in St. Louis (Knight-ADRC), and Emory University. Phenotypes and covariates (case/ control status, age, *APOE* genotypes, and education level) were not defined consistently across studies, and were re-categorized to be as consistent as possible (Supplementary Methods). Individuals under the age of 55 were excluded from the dataset. To minimize the effect of population stratification, only individuals of European ancestry were included, as determined by the first two genetic principal components. A total of 9139 participants were included (Table 1).

**Table 1.** Overview of study participants. Age and Education are presented as mean and standard deviation in years, sex as number of males, and *APOE* as count for 0, 1, and 2 alleles.

| Covariate | Cross-Validation |  | Holdout (ADNI3) |  |
| --- | --- | --- | --- | --- |
|  | Cases<br>(n = 3749, 41%) | Controls<br>(n = 5390, 59%) | Cases<br>(n = 59, 11%) | Controls<br>(n = 447, 89%) |
| Age | 78.0 (8.8) | 74.3 (8.7) | 73.0 (9.5) | 71.7 (6.2) |
| Sex | 1868 (49%) | 3165 (59%) | 22 (37%) | 283 (63%) |
| *APOE*-ε2 | 3470, 274, 5 | 4619, 744, 27 | 56, 3, 0 | 407, 40, 0 |
| *APOE*-ε4 | 1752, 1518, 479 | 3923, 1351, 116 | 17, 27, 15 | 286, 144, 17 |
| Education | 15.1 (3.1) | 15.9 (2.6) | 15.8 (2.8) | 16.8 (2.2) |

The ADNI3 dataset was held out as an independent validation set. After removing individuals related to or included in the main dataset, the ADNI3 data consisted of 316 individuals, assessed at multiple ages, for a total of 681 records. There were 28 unique cases and 238 unique controls. There were 77 unique instances of mild cognitive impairment (MCI), which were excluded from modeling. Participants were included at multiple ages if possible, to evaluate the effect of paragenic risk score on disease progression.

### 2.2. Data Collection

For data collected on the Emory University cohort, all research participants provide informed consent for blood and CSF collection and allowed clinical and biospecimen data to be repurposed under protocols approved by the Institutional Review Board of Emory University. A clinical diagnosis using standard clinical research criteria was assigned by a neurologist with subspecialty training in behavioral neurology. Blood and CSF were collected using a standardized approach from volunteers who were asked to fast at least 6 hours prior to collection. Genotyping was performed using the Affymetrix Precision Medicine Array using DNA extracted from the buffy coat by the Qiagen GenePure kit following the manufacturer’s recommended protocol.

### 2.3. Genotypes

Different genotyping chips were used across studies; therefore, genotypes from all studies were imputed to the Haplotype Reference Consortium (HRCr1.1) panel using the Michigan Imputation Server [14]. All files were prepared for imputation using the provided perl script (`HRC-1000G-check-bim.pl`). The imputed genotypes were filtered to biallelic SNPs with *Rsq* > 0.8 in all studies. SNPs with large differences in minor allele frequency (MAF) across studies or with potential strand flips were also removed. KING [15] was used to identify duplicate participants that were then removed from the dataset. Variants were then filtered to include those with *MAF* > 0.1 using PLINK.

### 2.4. Model Overview

To compare different modeling strategies and feature sources, we trained and evaluated several different models on our dataset. Throughout this work, we will use the following terminology to refer to different models trained to predict AD status:

1. *Baseline model:* a gradient boosting model trained on age, sex, and *APOE* genotype.
2. *PRS model:* a logistic regression model in which a PRS was computed and used as a feature along with the features of the baseline model.
3. *Epistatic model:* any model trained on mined epistatic features along with individual SNP markers and other covariates. We used two separate epistatic models trained on the same features, one using gradient boosting machines, and one using neural networks.
4. *Ensemble model:* any model trained on the predictions of other models.
5. *Paragenic model:* any ensemble model containing a PRS model and at least one epistatic model.

### 2.5. Epistatic Models

A feature engineering and association testing pipeline was run to select individual SNPs as well as interactions between SNPs to include in the individual epistatic models. The selected features, along with covariates (age, sex, *APOE*, education level, and the first 20 genomic principal components) were used to separately train and validate gradient boosting machine (XGBoost gradient boosting classifier algorithm [16]) and neural network (Neural Oblivious Decision Ensemble neural network (NODEnn) architecture [17]) models predicting case/control status. Feature selection and model building were performed using 10-fold nested cross-validation on the aggregated dataset. The same cross-validation fold partitions were used throughout.

#### 2.5.1 Individual SNP Selection

Individual SNPs were selected by linear mixed modeling association with case/control status using BOLT [18]. The participants were randomly partitioned into 10 crossvalidation folds. Related individuals were detected using KING [15] and assigned to the same fold for the mixed model association testing step, after which the maximum unrelated set for each family group was computed and retained. The resulting training sets (each consisting of nine folds) had a mean of 3374 (sd 18.9) cases and 4851 (sd 23.5) controls, and the test sets (one fold) had a mean of 375 (sd 18.9) cases and 539 (sd 23.5) controls. The top 50 SNPs ranked by log odds ratio were included as features in the modeling step.

#### 2.5.2 Epistatic Interaction Feature Engineering

Epistatic interaction terms were selected using multifactor dimensionality reduction (MDR) [19], a nonparametric approach that collapses the genotype combinations into high risk or low risk, then tests this new variable’s association with the phenotype using cross validation. The Crush-MDR algorithm [13] uses an evolutionary algorithm guided by expert knowledge to mine the space of SNP interactions. Candidate SNPs to include in the interaction mining were selected within each training set of unrelated individuals. To reduce the dataset to a size that could fit in memory for the epistatic feature pipeline, we used PLINK to remove SNPs in linkage disequilibrium (LD) to downsample to approximately 100,000 SNPs. We empirically chose a downsampling *r*^2^ > 0.11, which resulted in 98,903 SNPs. These SNPs were then run through an iterated version of the MultiSURF algorithm [20], and the top 10,000 SNPs associated with disease status were retained. The space of epistatic interaction terms using combinations of either two or three SNPs was mined using the Crush-MDR algorithm with multi-objective optimization and expert knowledge provided in the form of the number of shared pathways between each pair of SNPs as well as pairwise mutual information conditioned on case/control status. Shared pathways were computed using annotations from the Gene Ontology database [21, 22]. Each SNP was associated with the gene closest in distance to it, or containing it if there was such a gene. SNPs were considered to share a pathway if the associated genes shared a pathway as defined in the Gene Ontology database. Interaction terms were ranked by Pareto optimality with respect to balanced accuracy and mean cartesian entropy, and the top 100 were selected as features for downstream modeling. Interaction terms were represented for each individual as whether they had a low risk or high risk genotype combination. Genotype combinations that were not found in the training set were coded as missing.

#### 2.5.3 Gradient Boosting Model

The variants, epistatic terms, and covariates selected above were provided to XGBoost [16], a gradient boosting machines algorithm, as features. Within each cross-validation training fold, hyperparameters were tuned in an inner crossvalidation loop using Origin [23], a distributed implementation of the nondominated sorting genetic algorithm II (NSGA II) [24]. Origin was run on a cluster of Amazon Web Services spot instances.

#### 2.5.4 Neural Network Model

The NODEnn architecture does not support missing feature values; therefore, only variants and epistatic terms were included in the modeling as there was no way to meaningfully impute the covariates. Imputed genotype dosage values were used in place of allele counts for single SNPs to minimize missing values; any remaining missing values were imputed using k-nearest neighbors (k = 5) imputation on the training set. After imputation, the dosage and epistatic features were normalized to be between 0 and 1. The NODEnn model was constructed using the PyTorch implementation provided by the original authors [17]. Our network consisted of two blocks, each consisting of 1024 neural trees with depth = 6 and dimension = 3 and quasi-hyperbolic Adam [25] with the recommended hyperparameter settings of *ν*_0_ = 0.7, *ν*_1_ = 1.0, /3_0_ = 0.95 and *β*_1_ = 0.998 as the optimizer. The network was trained on an NVIDIA Titan RTX GPU. The NODEnn model was regularized using early stopping. On each fold, 10% of the training set was held out as a validation set; training was stopped when the model failed to improve after 5 epochs.

### 2.6. Calculation of PRS Model

Construction of a polygenic risk score (PRS) was performed according to the methodology of [26]. The same cross-validation folds as the epistatic model were used; the training sets were used as discovery sets in the PRS model and the test sets were used for validation. Standard quality control procedures were applied: only SNPs having MAF ≥ 0.01, Hardy–Weinberg equilibrium x^2^ test pvalue ≥ 1 × 10^*-*6^ and genotyping call rate ≥ 0.9 were included in the discovery set [27]. We removed individuals with genotype missingness ≥ 0.1 and randomly removed related individuals with a kinship coefficient cutoff of 0.125. We performed random linkage disequilibrium pruning and intelligent pruning with the --clump option in PLINK using *r*^2^ > 0.2 and a physical distancing threshold of 1Mb to be consistent with [26].

Markers were selected using p-value thresholds ranging from 0.05 to 1.0, and the polygenic risk score was calculated with effect sizes from the IGAP study as weights [28]. Performance of the PRS models were computed using logistic regression on disease risk against PRS, *APOE*-*ε*2, *ε*4, genotype, age, and sex using the StatsModels package [29]. The model with significance threshold p = 0.6 resulted in the PRS model with the highest AUC, and was used for comparative analysis.

### 2.7. Paragenic and Ensemble Models

To train the ensemble models, within-training-set predictions were computed on each fold for the XGBoost and NODEnn epistatic models as well as the PRS model. For each training fold, these predictions were provided as features to train an ensemble model by stacking [30] with logistic regression as a meta model using Scikit-Learn [31]. Predictions for the stacked model were then computed on the test set for each fold and used for final ensemble model evaluation. We also trained and evaluated an ensemble model for each of the various combinations of individual XGBoost, NODEnn, and PRS models. Figure 1 summarizes the modeling process.

**Figure 1.**
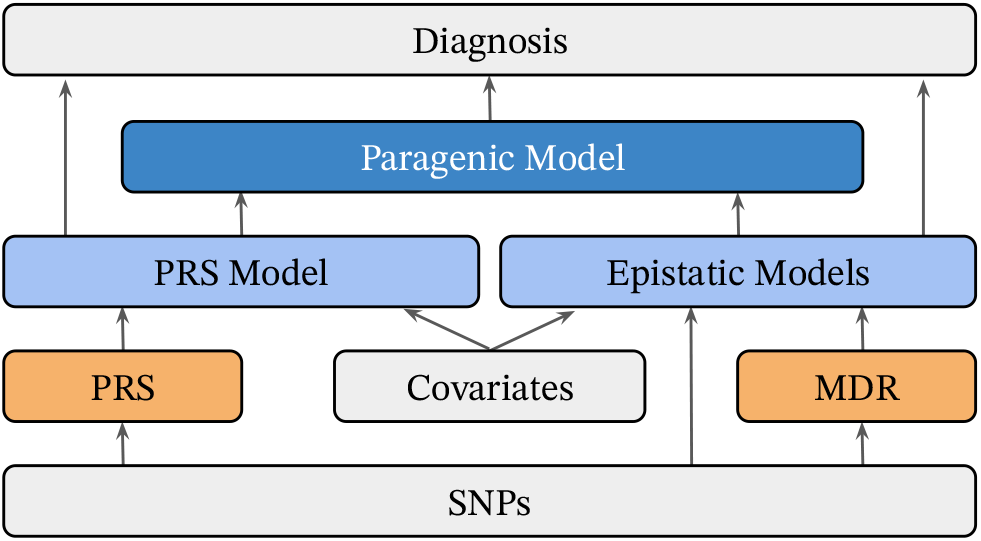
Schematic of mining and modeling procedure. Raw data is labeled in gray, derived features in orange, and models in blue.

## 3. Results

The paragenic ensemble models were compared to their individual component models as well as all combinations of the individual components. A baseline logistic regression model on age, sex, and number of *APOE*-*ε*2 and -*ε*4 alleles was included as well. Model efficacy was determined by area under the receiver operator characteristic curve (ROC AUC or just AUC), specificity and sensitivity. Models were evaluated both by cross-validation performance and in an independent holdout set of ADNI3 participants who were new to the model.

### 3.1. Model Performance

Models were first assessed using the test set predictions from 10-fold cross-validation. Model AUC was calculated for each test set (Table 2). Probability thresholds minimizing the difference between sensitivity and specificity were computed in the training sets for each fold and then applied to the test sets; standard deviations of sensitivity and specificity statistics were computed using bootstrap sampling.

**Table 2.** Comparison of individual models and ensembles on cross-validation, calculated as mean and 95% confidence intervals across folds

| Model | ROC AUC | Specificity | Sensitivity |
| --- | --- | --- | --- |
| Baseline (age + sex + APOE) | 0.749 (0.739, 0.759) | 0.735 (0.723, 0.747) | 0.641 (0.626, 0.657) |
| PRS | 0.796 (0.786, 0.805) | 0.757 (0.746, 0.769) | 0.698 (0.683, 0.712) |
| XGBoost | 0.804 (0.795, 0.813) | 0.707 (0.695, 0.719) | **0.746 (0.732, 0.759)** |
| NODEnn | 0.727 (0.716, 0.737) | 0.687 (0.675, 0.699) | 0.648 (0.632, 0.664) |
| PRS + XGBoost | 0.829 (0.820, 0.837) | 0.758 (0.745, 0.769) | 0.741 (0.726, 0.755) |
| XGBoost + NODEnn | 0.803 (0.794, 0.812) | 0.711 (0.699, 0.722) | 0.740 (0.725, 0.753) |
| PRS + NODEnn | 0.801 (0.792, 0.810) | 0.726 (0.714, 0.737) | 0.738 (0.724, 0.752) |
| PRS + XGBoost + NODEnn | **0.829 (0.821, 0.838)** | **0.761 (0.750, 0.772)** | 0.736 (0.722, 0.749) |

The paragenic model on PRS + XGBoost + NODEnn significantly outperformed all individual models in terms of AUC and specificity (DeLong test statistic *Z* = − 3.2555, p-value = 0.0006, McNemar test statistic for specificity *χ*^2^ = 553.0, p-value < 10^−10^ between the paragenic model and the model with the next highest AUC/specificity). The standard PRS model performed significantly better than the model ensembles in terms of specificity (*χ*^2^ = 553.0, p-value < 10^−10^), but at the cost of having a significantly lower sensitivity than the least performant paragenic model (PRS + NODEnn, *χ*^2^ = 88.0, p-value < 10^−16^).

Models were then trained on the entire dataset and predictions were made on the ADNI3 holdout set. Again, the minimized difference between sensitivity and specificity probability threshold was computed on the training set, and sensitivity and specificity standard deviations were computed via bootstrapping in the holdout set (Table 3). Comparison of ROC curves in the cross-validation and ADNI3 sets are shown in Figure 2.

**Table 3.**
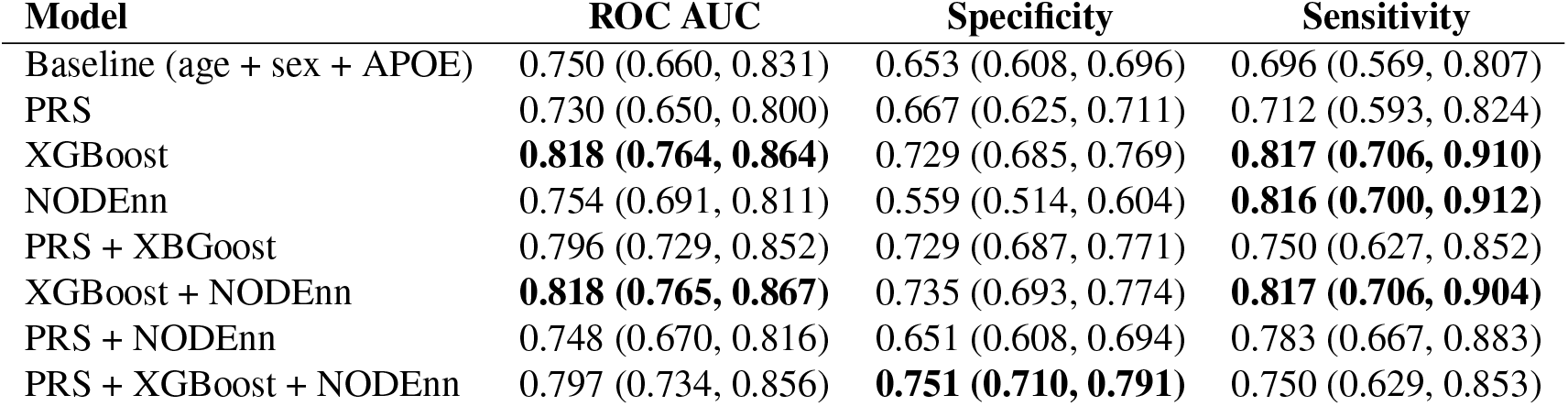
Comparison of individual models and ensembles on holdout dataset (ADNI3).

| Model | ROC AUC | Specificity | Sensitivity |
| --- | --- | --- | --- |
| Baseline (age + sex + APOE) | 0.750 (0.660, 0.831) | 0.653 (0.608, 0.696) | 0.696 (0.569, 0.807) |
| PRS | 0.730 (0.650, 0.800) | 0.667 (0.625, 0.711) | 0.712 (0.593, 0.824) |
| XGBoost | **0.818 (0.764, 0.864)** | 0.729 (0.685, 0.769) | **0.817 (0.706, 0.910)** |
| NODEnn | 0.754 (0.691, 0.811) | 0.559 (0.514, 0.604) | **0.816 (0.700, 0.912)** |
| PRS + XGBoost | 0.796 (0.729, 0.852) | 0.729 (0.687, 0.771) | 0.750 (0.627, 0.852) |
| XGBoost + NODEnn | **0.818 (0.765, 0.867)** | 0.735 (0.693, 0.774) | **0.817 (0.706, 0.904)** |
| PRS + NODEnn | 0.748 (0.670, 0.816) | 0.651 (0.608, 0.694) | 0.783 (0.667, 0.883) |
| PRS + XGBoost + NODEnn | 0.797 (0.734, 0.856) | **0.751 (0.710, 0.791)** | 0.750 (0.629, 0.853) |

**Figure 2.**
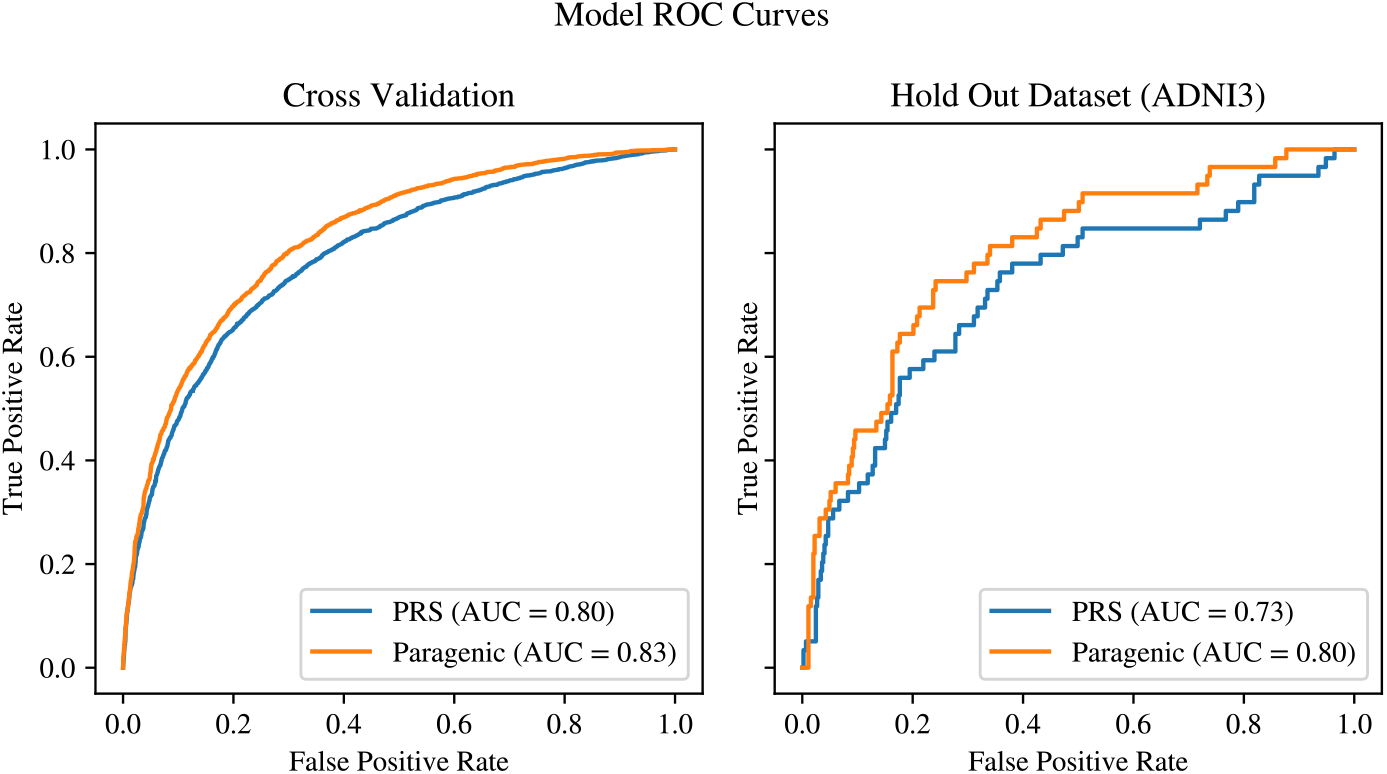
Comparison of ROC curves between PRS and Paragenic models on cross-validation and hold out data (ADNI3).

The XGBoost model outperformed all other models in the holdout set in terms of AUC (*Z* = −42.9, p-value < 10^−15^) and had significantly higher sensitivity over PRS (*χ*^2^ = 6.0, p-value = 0.0003). It had significantly higher specificity among models incorporating SNP or epistatic features (*χ*^2^ = 79.0, p-value = 4.5 ×10^−5^). The baseline model was the most specific (*χ*^2^ = 82.0, p-value = 0.0006); however it had extremely poor sensitivity and AUC and therefore is less performant overall. Interestingly, the ensemble models (even those including XGBoost) failed to outperform the component models on the holdout set; in particular, inclusion of PRS in an ensemble was generally detrimental to performance. Despite the XGBoost model performing well on the holdout set, this performance falls off significantly in terms of specificity and AUC when ensembled with PRS (*Z* = 42.9, p-value < 10^−15^, *χ*^2^ = 149.0, p-value < 10^−30^).

### 3.2. Model Performance by *APOE* Genotype

The full paragenic model (PRS + XGBoost + NODEnn) showed strong AUC performance within all *APOE* genotypes in the cross-validation dataset, significantly outperforming PRS within all *APOE* genotypes (Figure 3). The paragenic model performed consistently well within each stratum, staying within 4% points of the unstratified AUC for all genotypes except *ε*4/*ε*4. The PRS model largely performed much poorer within each stratum compared to the unstratified AUC, generally 6–7% lower with the exception of the *ε*2/*ε*4 stratum. Both models had AUCs on the *ε*2/*ε*4 stratum on par with their respective unstratified AUCs. The *ε*2/*ε*2 genotype was not well-represented in our data and so was discarded from this analysis.

**Figure 3.**
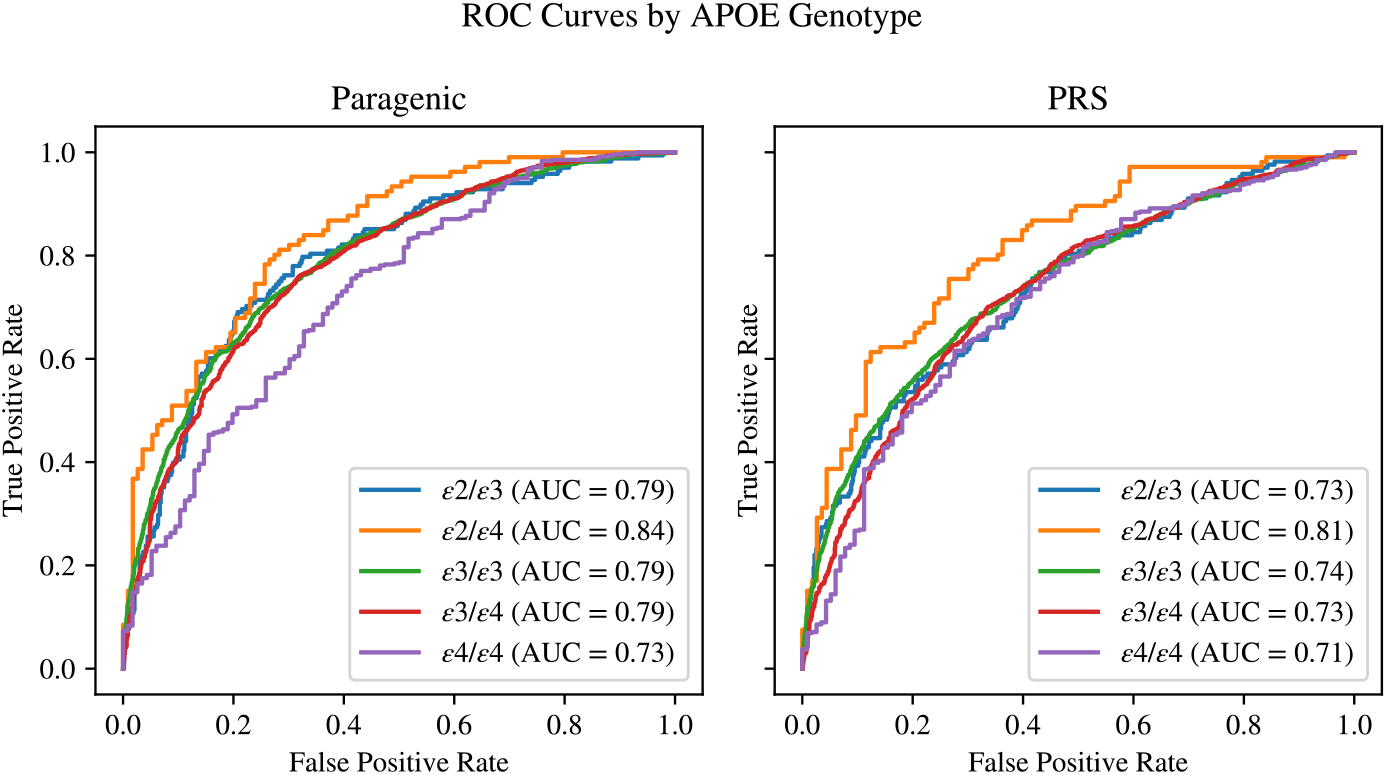
Comparison of ROC curves between PRS and Paragenic models on cross-validation and hold out data (ADNI3).

### 3.3. Risk Prediction by Age

To assess age-dependent risk, the Kaplan–Meier survival curve was estimated on the cross-validation dataset within each score quantile of the full paragenic model using the lifelines package in Python [32]. The resulting curve showed significant discrimination for LOAD risk at different ages (Figure 4). The logrank p-values between Q1 and Q2, Q2 and Q3, Q3 and Q4 were 0.0016, < 10^−14^, and < 10^−17^ respectively.

**Figure 4.**
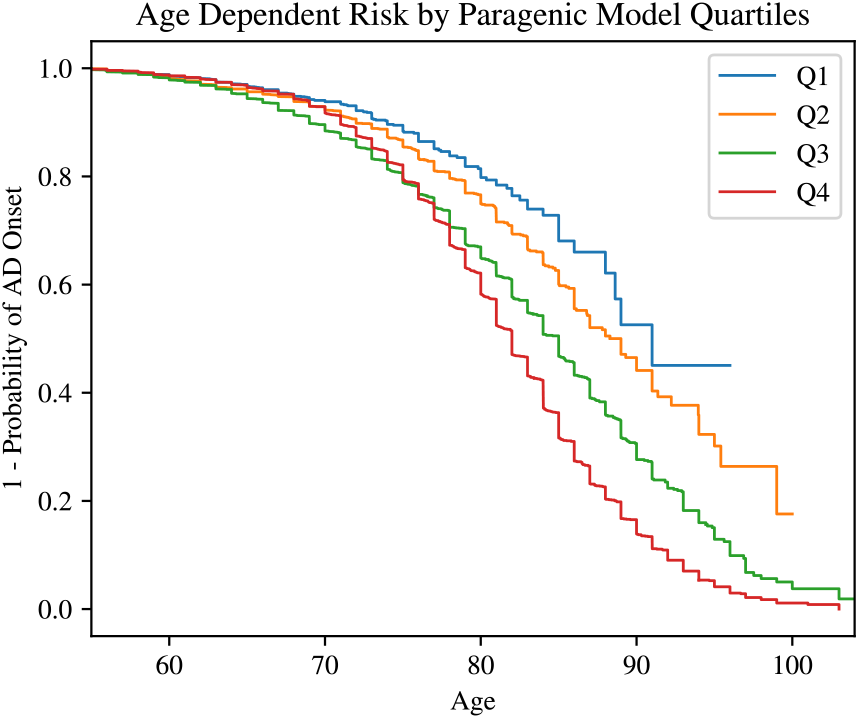
Comparison of ROC curves between PRS and Paragenic models on cross-validation and hold out data (ADNI3).

### 3.4. Clinical Utility

To assess our model for clinical utility, we analyzed the positive and negative predictive values (PPV and NPV) in Python. Following [26], we computed adjusted PPV and NPV values assuming varying LOAD population prevalences of 17% (overall lifetime risk) and 32% (risk for ages 85+ [33]). The results for cross-validation are presented in Table 4.

**Table 4.** Comparison of positive and negative predictive value of individual models and ensembles on cross-validation data.

| Model | Negative Predictive Value |  | Positive Predictive Value |  |
| --- | --- | --- | --- | --- |
|  | 17% prevalence | 32% prevalence | 17% prevalence | 32% prevalence |
| Baseline (age + sex + APOE) | 0.915 (0.911, 0.919) | 0.824 (0.817, 0.832) | 0.301 (0.288, 0.315) | 0.495 (0.476, 0.516) |
| PRS | 0.928 (0.925, 0.932) | 0.849 (0.842, 0.856) | 0.353 (0.338, 0.370) | 0.557 (0.535, 0.578) |
| XGBoost | 0.928 (0.925, 0.932) | 0.849 (0.842, 0.855) | 0.357 (0.341, 0.374) | 0.563 (0.543, 0.584) |
| NODEnn | 0.910 (0.906, 0.914) | 0.815 (0.807, 0.823) | 0.266 (0.254, 0.278) | 0.446 (0.429, 0.465) |
| PRS + XGBoost | **0.935 (0.932, 0.939)** | **0.863 (0.856, 0.869)** | **0.386 (0.369, 0.403)** | **0.593 (0.572, 0.615)** |
| XGBoost + NODEnn | 0.927 (0.923, 0.930) | 0.847 (0.839, 0.854) | 0.358 (0.342, 0.376) | 0.565 (0.544, 0.586) |
| PRS + NODEnn | 0.932 (0.929, 0.936) | 0.857 (0.850, 0.864) | 0.342 (0.327, 0.359) | 0.539 (0.519, 0.559) |
| PRS + XGBoost + NODEnn | **0.935 (0.932, 0.939)** | **0.863 (0.856, 0.869)** | **0.386 (0.370, 0.405)** | **0.594 (0.574, 0.616)** |

The full paragenic model and the paragenic XGBoost + PRS model had nearly identical predictive value, and both were significantly better than the next closest contender.

The predictive values on the holdout set are consistent with the analysis on the cross-validation dataset. The full paragenic model had the strongest PPV at both prevalences analyzed, but poorer NPV than the purely epistatic models (XGBoost and NODEnn) (Table 5).

**Table 5.** Comparison of positive and negative predictive value of individual models and ensembles on holdout data (ADNI3).

| Model | Negative Predictive Value |  | Positive Predictive Value |  |
| --- | --- | --- | --- | --- |
|  | 17% prevalence | 32% prevalence | 17% prevalence | 32% prevalence |
| Baseline (age + sex + APOE) | 0.913 (0.883, 0.944) | 0.820 (0.764, 0.878) | 0.274 (0.232, 0.324) | 0.455 (0.391, 0.535) |
| PRS | 0.919 (0.889, 0.949) | 0.831 (0.773, 0.894) | 0.286 (0.241, 0.337) | 0.471 (0.402, 0.550) |
| XGBoost | **0.950 (0.925, 0.975)** | **0.893 (0.840, 0.946)** | 0.341 (0.290, 0.402) | 0.524 (0.456, 0.608) |
| NODEnn | 0.937 (0.902, 0.969) | 0.866 (0.802, 0.929) | 0.188 (0.159, 0.222) | 0.319 (0.278, 0.370) |
| PRS + XGBoost | **0.933 (0.905, 0.960)** | **0.861 (0.808, 0.915)** | **0.354 (0.301, 0.415)** | **0.550 (0.479, 0.641)** |
| XGBoost + NODEnn | **0.951 (0.924, 0.975)** | **0.892 (0.840, 0.945)** | 0.347 (0.296, 0.405) | 0.532 (0.462, 0.616) |
| PRS + NODEnn | 0.935 (0.905, 0.964) | 0.863 (0.803, 0.920) | 0.263 (0.225, 0.312) | 0.427 (0.370, 0.493) |
| PRS + XGBoost + NODEnn | **0.935 (0.907, 0.961)** | **0.864 (0.810, 0.917)** | **0.383 (0.325, 0.452)** | **0.593 (0.511, 0.690)** |

## 4. Discussion

In this study, we built a genetic risk prediction model for LOAD using machine learning techniques with the goal of improving upon existing PRS models. Though PRS models have successfully captured the additive genetic components of disease, they do not capture more complex genetic structure such as epistatic interactions. We constructed a PRS model on our dataset and additionally mined the data for epistatic features to include with single SNPs in machine learning models. The final paragenic risk model, an ensemble of a logistic PRS model, a deep learning epistatic model (NODEnn), and a gradient boosted trees epistatic model (XBGoost), achieved an AUC of 0.829 in cross-fold validation. This performance is significantly higher than published PRS models (Figures 2, 3) and, while other machine learning models have reported comparable or higher AUCs [34], they performed feature engineering in-sample, which can lead to overestimation of expected performance on realworld (out-of-sample) data [35].

The XGBoost epistatic model and paragenic models outperformed PRS in terms of ROC AUC, sensitivity, and specificity both in cross-validation and in an independent holdout set. Ensembling PRS and epistatic models generally improved the modeling over the individual component models across all metrics. As expected, all types of models performed less well on the held-out dataset than in crossvalidation. The XGBoost model suffered the least from data drift, particularly in terms of AUC. Ensembling of epistatic models with PRS did not give the same improvement as was observed in the cross-validation analysis, likely because the

PRS model performed poorly on the holdout dataset in general. We suspect this is due to the well-known difficulties in applying PRS models trained on one dataset to another [36]. However, ensembling with a PRS model did improve model specificity and positive predictive value on the holdout set. This issue was not apparent in the cross-validation study, likely because the folds were partitioned without stratification by data source, resulting in test/train splits comprising similar populations. Moreover, the holdout dataset had a significantly smaller proportion of cases, leading to poorer performance of the paragenic models in terms of specificity and NPV.

Importantly, the paragenic model showed improved discriminative ability over PRS alone regardless of *APOE* genotype. Although the “4/”4 genotype is a strong single marker predictor of LOAD, only 9.6% of people with AD carry this genotype and the prevalence is heterogeneous among populations [37]. Thus, predicting AD risk even in the absence of “4 alleles, and conversely predicting which *ε*4/*ε*4 carriers will not develop AD, is necessary. Interestingly, the genotype within which the paragenic model had the lowest ROC AUC was *ε*4/*ε*4. This may be due to the lower number of participants in this group and could possibly be improved through modeling within *APOE* genotypes, or synthetically increasing the prevalence of *ε*4/*ε*4 through oversampling.

It should be noted that the goal of this study was high predictive accuracy rather than interpretability or to provide insights into the etiology of LOAD. As such, feature significance was not explored in depth. Markers included in the model may be informative in predicting disease without being the true causative factor.

To the best of our knowledge, the paragenic model has the highest AUC of genetic risk prediction model for clinically diagnosed LOAD to date and can potentially be used to identify individuals at high and low risk of developing disease for stratification in clinical trials as well as for personal use. Further improvements can likely be made through inclusion of environmental and lifestyle covariates [38]. We found that the differences across studies in data collection methods and completeness for these factors resulted in informative missingness, and thus we were not able to use them in modeling. Inclusion of these factors in a personal risk prediction test would allow users to see how lifestyle changes can reduce their risk of developing disease. Additionally, this study, like others, was conducted on individuals of European ancestry only and thus sheds no insight into the efficacy of machine learning over PRS, which does not translate across ancestries, in non-Europeans. Modeling on diverse populations is required in order to extend this risk prediction test to individuals of all ancestries.

## Data Availability

The data supporting the findings of this study are available on request from the corresponding author. The data are not publicly available due to privacy or ethical restrictions.

## 5. Acknowledgements

We acknowledge the generous donations made by research volunteers at the Goizueta Alzheimer’s Disease Research Center at Emory University.

The recruitment and clinical characterization of research participants at the Knight-ADRC at Washington University were supported by NIH P30AG066444 (JCM), P01AG03991 (JCM), and P01AG026276 (JCM). This work was supported by grants from the National Institutes of Health (R01AG044546 (CC), P01AG003991 (CC, JCM), RF1AG053303 (CC), RF1AG058501 (CC), U01AG058922 (CC), RF1AG074007 (YJS)), the Chan Zuckerberg Initiative (CZI), the Michael J. Fox Foundation (CC), the Department of Defense (LI-W81XWH2010849), and the Alzheimer’s Association Zenith Fellows Award (ZEN-22-848604, awarded to CC). This work was supported by access to equipment made possible by the Hope Center for Neurological Disorders, the NeuroGenomics and Informatics Center and the Departments of Neurology and Psychiatry at Washington University School of Medicine.

Data collection and sharing for this project was funded by the Alzheimer’s Disease Neuroimaging Initiative (ADNI) (National Institutes of Health Grant U01 AG024904) and DOD ADNI (Department of Defense award number W81XWH-12-2-0012). ADNI is funded by the National Institute on Aging, the National Institute of Biomedical Imaging and Bioengineering, and through generous contributions from the following: AbbVie, Alzheimer’s Association; Alzheimer’s Drug Discovery Foundation; Araclon Biotech; BioClinica, Inc.; Biogen; Bristol-Myers Squibb Company; CereSpir, Inc.; Cogstate; Eisai Inc.; Elan Pharmaceuticals, Inc.; Eli Lilly and Company; EuroImmun; F. Hoffmann-La Roche Ltd and its affiliated company Genentech, Inc.; Fujirebio; GE Healthcare; IXICO Ltd.; Janssen Alzheimer Immunotherapy Research & Development, LLC.; Johnson & Johnson Pharmaceutical Research & Development LLC.; Lumosity; Lundbeck; Merck & Co., Inc.; Meso Scale Diagnostics, LLC.; NeuroRx Research; Neurotrack Technologies; Novartis Pharmaceuticals Corporation; Pfizer Inc.; Piramal Imaging; Servier; Takeda Pharmaceutical Company; and Transition Therapeutics. The Canadian Institutes of Health Research is providing funds to support ADNI clinical sites in Canada. Private sector contributions are facilitated by the Foundation for the National Institutes of Health (www.fnih.org). The grantee organization is the Northern California Institute for Research and Education, and the study is coordinated by the Alzheimer’s Therapeutic Research Institute at the University of Southern California. ADNI data are disseminated by the Laboratory for Neuro Imaging at the University of Southern California. The NACC database is funded by NIA/NIH Grant U24 AG072122 and the NACC data are contributed by the NIA-funded ADRCs: P30 AG062429 (PI James Brewer, MD, PhD), P30 AG066468 (PI Oscar Lopez, MD), P30 AG062421 (PI Bradley Hyman, MD, PhD), P30 AG066509 (PI Thomas Grabowski, MD), P30 AG066514 (PI Mary Sano, PhD), P30 AG066530 (PI Helena Chui, MD), P30 AG066507 (PI Marilyn Albert, PhD), P30 AG066444 (PI John Morris, MD), P30 AG066518 (PI Jeffrey Kaye, MD), P30 AG066512 (PI Thomas Wisniewski, MD), P30 AG066462 (PI Scott Small, MD), P30 AG072979 (PI David Wolk, MD), P30 AG072972 (PI Charles DeCarli, MD), P30 AG072976 (PI Andrew Saykin, PsyD), P30 AG072975 (PI David Bennett, MD), P30 AG072978 (PI Neil Kowall, MD), P30 AG072977 (PI Robert Vassar, PhD), P30 AG066519 (PI Frank LaFerla, PhD), P30 AG062677 (PI Ronald Petersen, MD, PhD), P30 AG079280 (PI Eric Reiman, MD), P30 AG062422 (PI Gil Rabinovici, MD), P30 AG066511 (PI Allan Levey, MD, PhD), P30 AG072946 (PI Linda Van Eldik, PhD), P30 AG062715 (PI Sanjay Asthana, MD, FRCP), P30 AG072973 (PI Russell Swerdlow, MD), P30 AG066506 (PI Todd Golde, MD, PhD), P30 AG066508 (PI Stephen Strittmatter, MD, PhD), P30 AG066515 (PI Victor Henderson, MD, MS), P30 AG072947 (PI Suzanne Craft, PhD), P30 AG072931 (PI Henry Paulson, MD, PhD), P30 AG066546 (PI Sudha Seshadri, MD), P20 AG068024 (PI Erik Roberson, MD, PhD), P20 AG068053 (PI Justin Miller, PhD), P20 AG068077 (PI Gary Rosenberg, MD), P20 AG068082 (PI Angela Jefferson, PhD), P30 AG072958 (PI Heather Whitson, MD), P30 AG072959 (PI James Leverenz, MD). The Alzheimer’s Disease Genetic Consortium (ADGC), is funded by NIA/NIH Grant U01 AG032984. From the Framingham Heart Study of the National Heart Lung and Blood Institute of the National Institutes of Health and Boston University School of Medicine. This project has been funded in whole or in part with Federal funds from the National Heart, Lung, and Blood Institute, National Institutes of Health, Department of Health and Human Services, under Contract No. 75N92019D00031.

## Footnotes

https://neurogenomics.wustl.edu/

