## Supplementary Methods for "Epistatic Features and Machine Learning Improve Alzheimer’s Risk Prediction Over Polygenic Risk Scores"

### A. Supplementary Methods

#### A.1. Datasets

##### A.1.1 ADNI

SNP genotype and phenotype data from ADNI1, ADNI2/GO, WGS, and ADNI3 cohorts were downloaded from the ADNI LONI database. The ADNI1 project, the ADNIGO/2 project, and the whole genome sequencing (WGS) project were included in modeling. Subjects were genotyped using Illumina Human610-Quad BeadChip, Illumina HumanOmniExpress BeadChip, and Illumina Omni 2.5M Chip respectively. The ADNI3 project, genotyped with the Illumina Infinium Global Screening Array v2 Chip, was used as an independent validation set.

##### A.1.2 Framingham Heart Study

Genotypes and phenotypes from the Framingham Heart Study (FHS) were downloaded from dbGAP (phs000007). The Framingham SHARe (phs000342.v18.p11), Framingham CARe (phs000282.v19.p11), and Framingham CHARGE-WGS (phs000651.v10.p11) sub-studies were included.

##### A.1.3 NACC/ADGC

Genotype data from the Alzheimer's Disease Genetics Consortium (ADGC) cohorts ADC1-3 were obtained from dbGAP (phs000372.v1.p1) and cohorts ADC4-7 were obtained from NIAGADS (NG00068, NG00069, NG00070, NG00071). Phenotype information for these subjects was obtained from the National Alzheimer's Coordinating Center (NACC). Subjects from ADC1 and ADC2 were genotyped on the Illumina Human660-Quad BeadChip, ADC3-6 were genotyped on the Illumina HumanOmniExpress BeadChip, and ADC7 was genotyped on the Illumina OmniExpress Exome chip.

##### A.1.4 Knight-ADRC

The recruited individuals were evaluated by Clinical Core personnel of the Knight ADRC. Research participants at the Knight-ADRC undergo longitudinal cognitive, neuropsychologic, imaging, and biomarker assessments including Clinical Dementia Rating (CDR). Among individuals with CSF and plasma data, AD cases corresponded to those with a diagnosis of dementia of the Alzheimer's type (DAT) using criteria equivalent to the National Institute of Neurological and Communication Disorders and Stroke. Alzheimer's Disease and Related Disorders Association for probable AD [39] and AD severity was determined using the Clinical Dementia Rating (CDR) [40] at the time of lumbar puncture (for CSF samples) or blood draw (for plasma samples). Age at onset was defined as the date the participant started showing cognitive impairment and was determined based on the longitudinal clinical assessment, including CDR and the semi-structured interview of the participant and a reliable informant or collateral source. Controls received the same assessment as the cases but were non-demented (CDR = 0).

Genotyping data come from several different rounds of genotyping on Illumina platforms. Stringent quality thresholds were applied to the genotype data for each platform separately. SNPs were kept if they met the following criteria: i) had a genotyping rate $\geq 98\%$; ii) had a MAF $\geq 0.3\%$; and iii) were in Hardy–Weinberg equilibrium (HWE) (p-value $> 10^{-6}$). After removing low quality SNPs and individuals, genotype imputation was performed using the Impute2 program with haplotypes derived from the 1,000 Genomes Project (released June 2012). Genotype imputation was performed separately based on the genotype platform used. SNPs were removed if they failed any of the following criteria: i) an impute2 info-score quality of less than 0.3; ii) a MAF ¡ 2%; or iii) out of HWE. After Imputation and QC, the different imputed plink files were merged.

To determine relatedness, Z0 and Z1 from IBD analysis for all individuals were plotted. Individuals which fell outside of the selected range ($Z0 \geq 0.65$, $Z1 \leq 0.4$) were considered relatives or duplicates. A single individual from each relative/duplicate pair with lowest call rate was removed. Finally, this analysis only used data from subjects of a European genetic background; genetic background for all individuals was determined by plotting the first two principal components (PCs) and identifying the European cluster.

##### A.1.5 Emory

All research participants provided informed consent for blood and CSF collection and allowed clinical and biospecimen data to be repurposed under protocols approved by the Institutional Review Board of Emory University. A clinical diagnosis using standard clinical research criteria was assigned by a neurologist with subspecialty training in behavioral neurology. Blood and CSF were collected using a standardized approach from volunteers who were asked to fast at least 6 hours prior to collection. Genotyping was performed using the Affymetrix Precision Medicine Array using DNA extracted from the buffy coat by the Qiagen GenePure kit following the manufacturer's recommended protocol.

http://adni.loni.usc.edu/

#### A.2. Phenotype Definitions

FHS was the only study that was not specifically conducted on Alzheimer's patients. The broader definition of dementia included in the FHS cases may have introduced some heterogeneity in the Case category. Phenotypes were defined per cohort as follows.

##### A.2.1 ADNI

Cases were defined by *ADNIMERGE DX = Dementia* and controls by *ADNIMERGE DX = CN*. Age was defined by the *AGE* field, and education by *ADNIMERGE Education*.

##### A.2.2 FHS

Cases were defined by *Cognitive Impairment (pht004368) demrv103* $> 0$ and $\neq 9$ OR *Dementia Flag Based on Neurology Exam (pht000690)* $> 1$ OR *Dementia Flag Based on Neuropsychological Test Battery (pht000691)* $> 1$. Controls defined by *Cognitive Impairment (pht004368) demrv103* = 0 OR *Dementia Flag Based on Neurology Exam (pht000690)* = 0 OR *Dementia Flag Based on Neuropsychological Test Battery (pht000691)* = 0. Age defined by *Cognitive Impairment (pht004368) min(dxmilddemdate, dxmoddemdate, dxsevdemdate, earlydemdate)* OR *cogimponsdate* if NA OR *review_date* if still NA. Education was defined as *Neuropsychological Battery (pht004374) education_b1* translated into years as $0 \to 0, 1 \to 3, 2 \to 6, 3 \to 8, 4 \to 10, 5 \to 12, 6 \to 14, 7 \to 16, 8 \to 18, 9 \to \text{NA}, 10 \to 15$ OR if *education_b1* not available, *education_b2 + 7* and capped at 20.

##### A.2.3 NACC/ADGC

Cases were defined by *NACCUDSD = 4* and controls by *NACCUDSD = 1*. Age was defined by *NACCAGE*, and education by *Education* capped at 20.

##### A.2.4 Knight-ADRC

Cases were defined by *CDR* $> 0.5$ and controls by *CDR = 0*. Age was defined by the *Age* field, and education by the *Education* field.

##### A.2.5 Emory

Cases were defined by *Dx = 1* and controls by *Dx = 0*. Age was defined by *AgeAtDiagnosis*, and education was marked NA.

#### A.3. *APOE* Covariates

*APOE* covariates were coded as the number of $\varepsilon 2$, $\varepsilon 3$, $\varepsilon 4$ alleles, as well as genotype: $0 = \varepsilon 2/\varepsilon 2$, $1 = \varepsilon 2/\varepsilon 3$, $2 = \varepsilon 3/\varepsilon 3$, $3 = \varepsilon 2/\varepsilon 4$, $4 = \varepsilon 3/\varepsilon 4$, $5 = \varepsilon 4/\varepsilon 4$. Some datasets contained explicit *APOE* covariates; if those conflicted with the genotype, they were set to missing.

#### A.4. Principal Components

Genotypes from 2,504 subjects from Phase 3 of the 1000 Genomes Project [41] were filtered to those SNPs that overlapped with the imputed dataset. AT/CG SNPs were removed, along with SNPs that had inconsistent alleles with the imputed data. The SNPs were then filtered using Plink `--indep-pairwise` with an LD threshold of $r^2 = 0.1$, a window size of 1 Mb, and a step size of 50 SNPs, leaving 42,135 SNPs. FlashPCA [42] was used to calculate the means, standard deviations, and loadings for the first 20 PCs. The imputed genotypes were then projected onto these PCs. Projecting PCs in this way ensures that consistent PC values can be obtained for new subjects.
